# SARS-CoV-2 risk taxation model and validation based on large scale Dutch test-events

**DOI:** 10.1101/2022.01.10.21268254

**Authors:** Bas Kolen, Laurens Znidarsic, Andreas Voss, Simon Donders, Iris Kamphorst, Maarten van Rijn, Dimitri Bonthuis, Merit Cloquet, Maarten Schram, Rutger Scharloo, Tim Boersma, Tim Stobernack, Pieter van Gelder

## Abstract

In response to the outbreak of SARS-CoV-2 many governments decided in 2020 to impose lockdowns on societies. Although the package of measures which constitute such lockdowns differs between countries, it is a general rule that contacts between people, and especially in large groups of people, are avoided or prohibited. The main reasoning behind these measures is preventing that healthcare systems become overloaded. As of 2021 vaccines against SARS-CoV-are available, but these do not guarantee 100% risk reduction and it will take a while for the world to reach a sufficient immune status. This raises the question whether and under which conditions events like theater shows, conferences, professional sports events, concerts and festivals can be organized. The current paper presents a COVID-19 Risk taxation method for (large scale) events. This method can be applied to events to define an alternative package of measures replacing generic social distancing.

## 1. Introduction

In response to the SARS-CoV-2 pandemic many governments implemented measures to reduce the risk for infections. Social distancing to reduce the number of contacts is the main measure to reduce the transmission of SARS-CoV-2 (*1*). To support social distancing, governments all over the world have taken measures, resulting in various types of (partial) lockdowns to reduce the number of contacts between people and limit gathering in groups. At the same time, Dutch event-organizers affirmed that organizing events on the basis of social distancing would be economically detrimental.

### 1.1 Research question

The aim of this research was to develop a model to determine the risk for infection at events during the pandemic, and the effectiveness of alternative measures instead of generic social distancing at these events. During test events we evaluated several measures and determined how the risk for infection at these events was compared to the average risk of infection during a lockdown or the risk at different locations. In this study we distinguish between four types of events:

- Type I: Indoor, passive (theater show or conference),
- Type II: Indoor, active (concert or dance events),
- Type III: Outdoor, active (public sports events),
- Type IV: Outdoor, active festival (festivals).

Regular contact matrices distinguish between different categories for locations and age. The most recent contact matrix for The Netherlands distinguishes between home, school, work and other as classes for locations, based on a pre pandemic situation (2). In these contact matrices also classes for age are used. Events are part of the class ‘other’. This class ‘other’ contains multiple locations which might contribute very differently to the risk for infections. Standard social contact data (2, 3) also do not apply because of governmental measures and personal behavior changes. We have therefore gathered actual data regarding contacts and settings representing peoples’ whereabouts during the pandemic, as well as statistics regarding the epidemiologic situation during the pandemic; they have been used to develop a causal risk taxation model.

In this study we have developed a causal model to describe the risk for infections at these events. Causal modelling forms an alternative for physical or biological models, and (as such) can support the insight into the interdependencies between the constituent parts of complex systems as these events (2). A causal risk model is developed based on available data of infections and contacts among people during the pandemic. This risk-model determines the risk for infection per hour at events depending on the circumstances of the event and the contacts at these events given the epidemiologic situation at that moment.

### 1.2 The infection risk per hour

Most of the available literature estimates the reproduction number R_0_ of Severe acute respiratory syndrome coronavirus 2 (SARS-CoV-2) is between 2-4 (*5, 6*). For Western Europe the R_0_ is estimated at 2.2 (*7*) and the Dutch National Institute for Public Health and the Environment (RIVM) estimated a range between 2 and 3 (*8*). The probability of an infected person infecting another person peaks during the first week of illness, after which this probability drops significantly (*9*). Based on data from Wuhan the incubation time was on average 5.2 days and at least 4 days (*10*). Later studies based on more data indicated that the average incubation time ranged between 5.2 and 6.65 days and could be up to 14 days (*12*). We decided to express the probability of infection during the period of exposure during an event as the probability per hour. This risk can be compared to the infection risk outside these events during the pandemic. When R_0_ is 2.2, people are infectious for 7 days and a prevalence of 0,77% in the Netherlands this means that the probability that a person infects another person is 1.04 10^−4^ per hour

The number of positive SARS-CoV-2 tests in the Netherlands are reported by the RIVM on a weekly basis (13). These reports include the expected location where people had been infected. Examples of these locations are homes, work, home while having visitors (friends or family), leisure, schools, catering industry, elderly houses and events. The measures taken by the government strongly influence the number of contacts at these locations. Although of all location types most people were infected at home, the probability per hour for a person of being infected at this type of location was relatively small compared to other types.

We conducted a survey te estimate the time people spend a several location and number of contacts at these locations. The individual probability per hour of being infected for each location can be compared when the absolute numbers of infections are corrected for the duration of the stay at a location and the number of people at these locations. These figures can be used as benchmarks when deciding whether to have events.

## 2. Methodology SARS-CoV-2 Risk taxation Model

### 2.1 Methodology

In this study a SARS-CoV-2 risk taxation model was developed based on data sets collected by the National Institute for Public Health and the Environment (RIVM) and Municipal health services (GGD) collected data sets during the SARS-CoV-2 pandemic. The method resulted in the average risk of infection at the event *i* per hour:

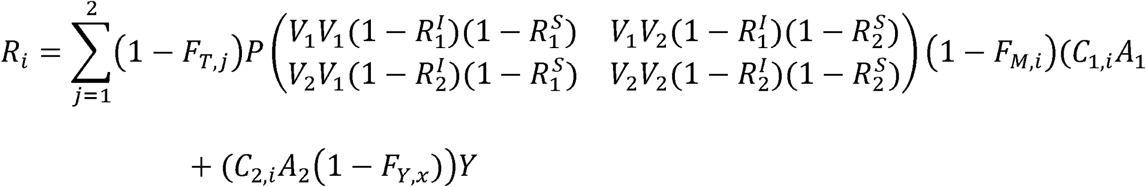

In which:

- The actual prevalence *p* describes the proportion of infectious people in the Netherlands.
- Contacts at the event *i*. The complete role of virus-laden droplets and aerosols transmission is poorly understood (*11*). We distinguish between two contact-classes which are most significant:*C*_1_ is the number of contacts per hour within 1,5m (droplets) and *C*_2_ the number of contacts per hour within 10m (aerosols). Smart logistics and crowd control at an event reduce contacts and avoid gathering of large groups of people. Therefore, specific data were collected at test events. Because the duration of events is limited the latency period is shorter than the duration of stay at each location.
- We consider two types of individuals *j* who can attend an event and for which different test regimes can apply. *j* = 1 corresponds to unvaccinated individuals without a documented infection,*j* = 2 corresponds to vaccinated individuals, or people with a documented infection. *V*_*j*_ is the proportion of the people in each group. The combination always equals 100%. At each event we assume an homogeneous mixing of all people.
- Testing prior to an event, factor *F*_*T,j*_ describes the proportion of people that cannot attend the event because of testing in a certain window prior to the event.
- Vaccination and earlier infections result in a level of immunity and reduces the infectiousness *I* and susceptibility *S* of an individual. 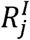 and 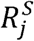 the relative infectiousness and susceptibility for a type *j* individual related to a naive person. For the test events used to validate the model all people were considered as naïve (which means that they do not have any immunity), during the test events nobody was vaccinated and a limited amount of people had had an infection.
- *F*_*Y,x*_.*x is* the percentage of reduction of aerosols. When *x* = 0 locations are ventilated according to building codes, *x* = 1 means very well ventilated and *x* = 2 is outdoor.
- Personal protection measures such as mouth-nose masks. This reduces the risk with factor *F*_*M*_.
- *Y* is a multiplier for variants which are more infectious than the Alfa-variant which was dominant during the data collection.

### 2.2 Transmission coefficients A1 and A2

The parameters *A*_1_ *and A*_2_ are transmission-coefficients for exposure to droplets and aerosols. We distinguished between two periods: 15 September – 13 October 2020 and 14 October - 15 December 2020. On 13 October additional measures were implemented by the Dutch Government, on the 15^th^ of December 2020 a new lockdown was implemented. For these two periods we collected contact data of persons by a questionnaire and combined with statistical data (14). We applied linear regression via Least-squares Minimization (as most common and proven approach for linear models) on a dataset of infections per location, duration and contact per location. We used data for the locations home, work, visitors at home and leisure. For the other locations the amount of available data was not sufficient. In the questionnaire we asked people after:

- The time spent at a certain location.
- The number of persons in a range of 10m.
- The number of contacts within a distance of < 0,5 m, between 0,5 and 1,5 and between 1,5 and 2,0 m for less than a minute, between 1 and 15 minutes and more than 15 minutes.
- The proportion of the time which was spend indoor, indoor and well-ventilated or outdoor given a type of location.

For each category of location, we assumed an average duration (30 seconds for the class < 1 minute, 8 minutes for contacts within 1 and 15 minutes and 30 minutes for contacts more than 15 minutes. For all indoor locations we assumed standard ventilated locations *F*_*y*,0_ = 0, for well ventilated indoor *F*_*y*,1_ =0.5, for outdoor events *F*_*y*,2_ = 1 All people were considered to be naïve, and testing was not available in this period.

This resulted in a probability per hour that a person will be infected given a contagious person in a distance of 1.5m (*A*_1_ = 6.376 10 ^-3^ per hour) or 10m (*A*_2_ =0.986 10 ^-3^ per hour).

Table 3 shows sensitivity analyses that the impact of other choices in the distances of 1,5m and 10m or for *F*_*y*_ is limited (given a prevalence of 0.5%). For the reference situation *C*_1_ is 5 contacts per hour for the low contact event and 12.5 for the high contact event. *C*_2_ is 10 contacts for the low contact event and 30 for the high contact event. The sensitivity analysis shows that the impact on the results sits within a bandwidth of a factor 0.5.

**Table 1:**
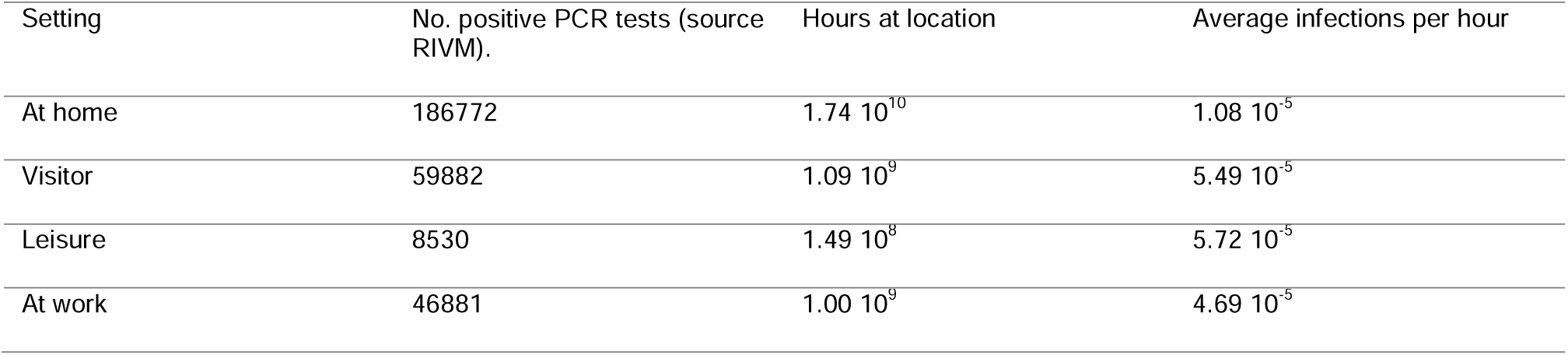
Location and infection data used for the least-squares regression of A1 and A2 based on RIVM and the Survey.

**Table 2:**
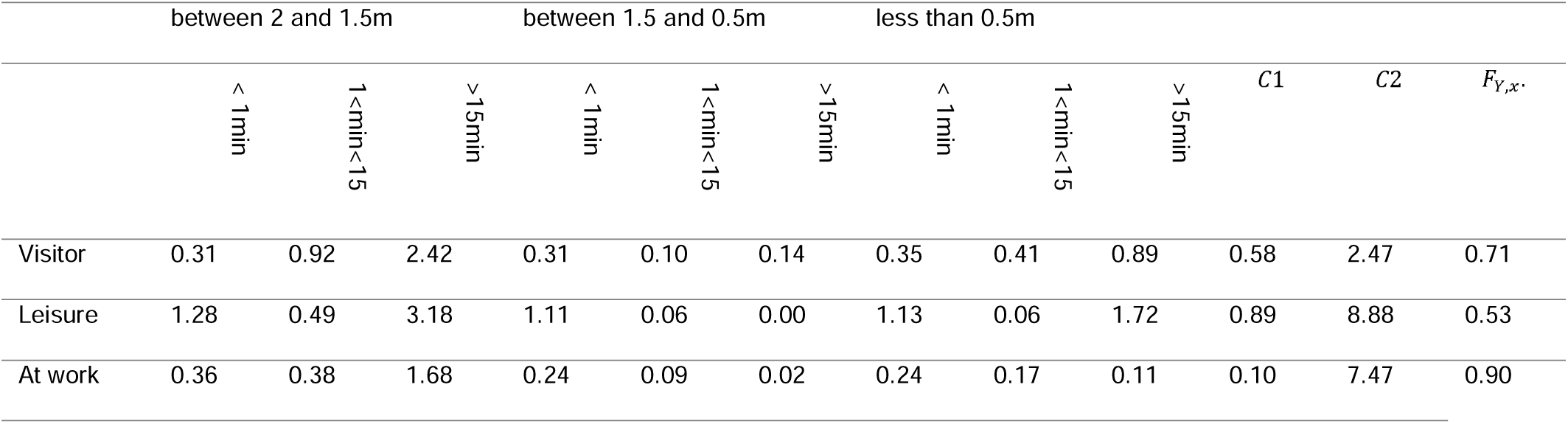
Contact data used for the least-squares regression of A1 and A2 based on RIVM and survey data on the number of contacts/hour within a certain contact category, per setting.

**Table 3:**
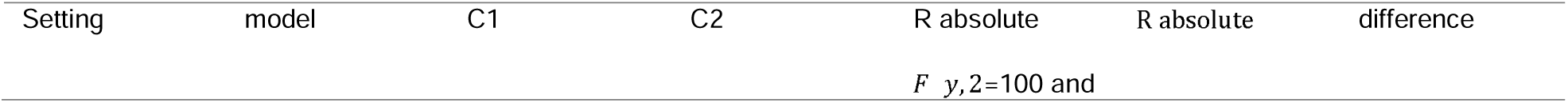
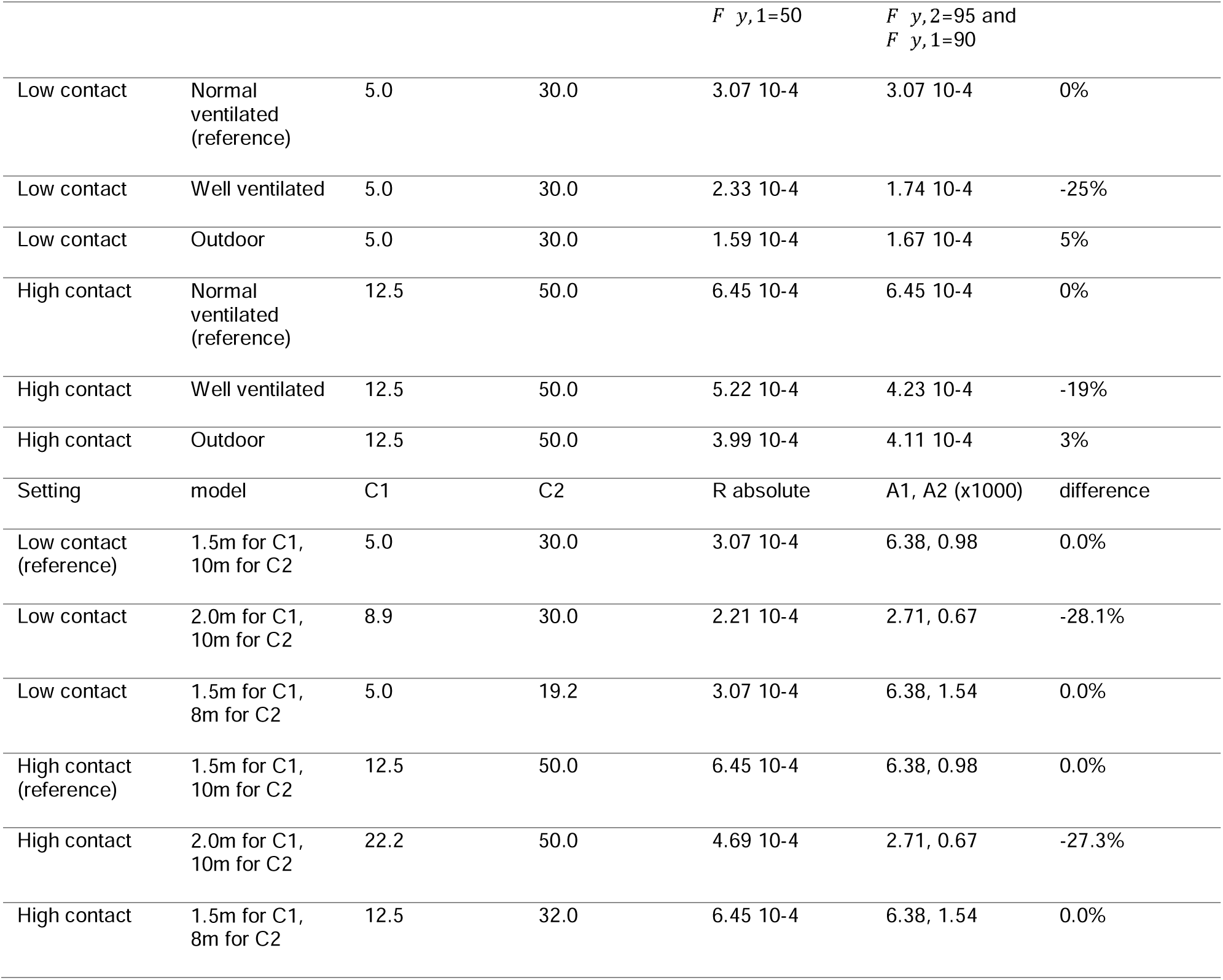
Sensitivity analyses for ventilation and distance for contact classes. F_V_, F_T_ and F_M_ are all 0

| Setting | model | C1 | C2 | R absolute | R absolute | difference |
| --- | --- | --- | --- | --- | --- | --- |
| | | | | $F_{y,2}=100$ and | | |

| | | | | $F_{y,1=50}$ | $F_{y,2=95}$ and<br>$F_{y,1=90}$ | |
| --- | --- | --- | --- | --- | --- | --- |
| Low contact | Normal ventilated (reference) | 5.0 | 30.0 | 3.07 10-4 | 3.07 10-4 | 0% |
| Low contact | Well ventilated | 5.0 | 30.0 | 2.33 10-4 | 1.74 10-4 | -25% |
| Low contact | Outdoor | 5.0 | 30.0 | 1.59 10-4 | 1.67 10-4 | 5% |
| High contact | Normal ventilated (reference) | 12.5 | 50.0 | 6.45 10-4 | 6.45 10-4 | 0% |
| High contact | Well ventilated | 12.5 | 50.0 | 5.22 10-4 | 4.23 10-4 | -19% |
| High contact | Outdoor | 12.5 | 50.0 | 3.99 10-4 | 4.11 10-4 | 3% |
| Setting | model | C1 | C2 | R absolute | A1, A2 (x1000) | difference |
| Low contact (reference) | 1.5m for C1, 10m for C2 | 5.0 | 30.0 | 3.07 10-4 | 6.38, 0.98 | 0.0% |
| Low contact | 2.0m for C1, 10m for C2 | 8.9 | 30.0 | 2.21 10-4 | 2.71, 0.67 | -28.1% |
| Low contact | 1.5m for C1, 8m for C2 | 5.0 | 19.2 | 3.07 10-4 | 6.38, 1.54 | 0.0% |
| High contact (reference) | 1.5m for C1, 10m for C2 | 12.5 | 50.0 | 6.45 10-4 | 6.38, 0.98 | 0.0% |
| High contact | 2.0m for C1, 10m for C2 | 22.2 | 50.0 | 4.69 10-4 | 2.71, 0.67 | -27.3% |
| High contact | 1.5m for C1, 8m for C2 | 12.5 | 32.0 | 6.45 10-4 | 6.38, 1.54 | 0.0% |

### 2.2 Total number of infections at an event

The expected number of infections at an event *S* are the combination the product of *R*_*i*_ and the duration (in hours) of the event *t* and the number of people at the event *N* :

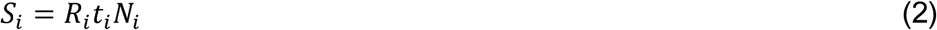

## 3. Validation of the model

The model determines an average risk for infection. The data concerning SARS-CoV-2 infections gathered at the test events can be used to validate the model. During these test vaccinations were not yet available, and only a small amount of people had earlier infections. Therefore all people were considered to be naïve, and all people who attended an event had to be tested (*V*_1_ = 100%). The model outcome, an average number of infections, is based on a skewed probability distribution. For example, consider a large-scale event where 1.000 people will join the event given a prevalence of 0.75%. Without pre-testing 7.5 infectious persons would have attended the event, when *F*_*T,j*_ on average 0.38 person would have been infectious at the event and could infect others. In the model we use the average number of contacts, there will be a probability distribution about these contacts some people will have many close contacts and others will see only a few people. Because of these uncertainties it is expected that many events will be organized that will result in no or limited number of infections, and a few events will result in many infections.

A model validation would need a large dataset during the SARS-CoV-2 pandemic. This large dataset is expected to cover the skewed probability distribution including events. Such a database, however, is not available. Data from media and literature of superspreader events are biased as these always attract more attention. For loss of life modelling for natural hazards the limited availability of data also leads to difficulties for conducting model validation. For example, the loss of life models for river and storm surge flooding as used in the Netherlands are based on the 1953 flood in Sealand and the flood caused by hurricane Katrina in 2005 in the US (*15, 16*). However, despite the limited validation, the model is still used to define the safety standards for Dutch levees which implies an investment program of multiple billions of euros (*17*). Although a perfect validation is not attainable, the available data can be used for a first validation of the model.

### 2.3.1 Internal validation: reproduction of infections at different settings

First, the performance of the model can be checked by the reproduction of infections for the settings at work, visitors and leisure time. Given the value for *A*_1_ and *A*_2_, the prevalence as published by the RIVM and using data from the questionnaire for the type of locations ‘visitors at home’, ‘work’ and ‘leisure’ to define *C*_*1*_, *C*_*2*_, *F*_*Y,x*_ we estimated the number of infections by the model for these locations. These model results are compared with the measured number of infection at these location by RIVM and regional health services (see figure 1). The infections ‘at work’ are overestimated in the model, while infections in the setting of ‘at home with visitors’ are underestimated. An explanation can be that people received more visitors than they admitted to in the survey or than allowed within the prevailing COVID-19-rules (two visitors per day until mid-October, and one per day afterwards). Overall, we concluded that the outcomes support the results of the model. Room for improvement is available if more contact data is available.

**Figure 1:**
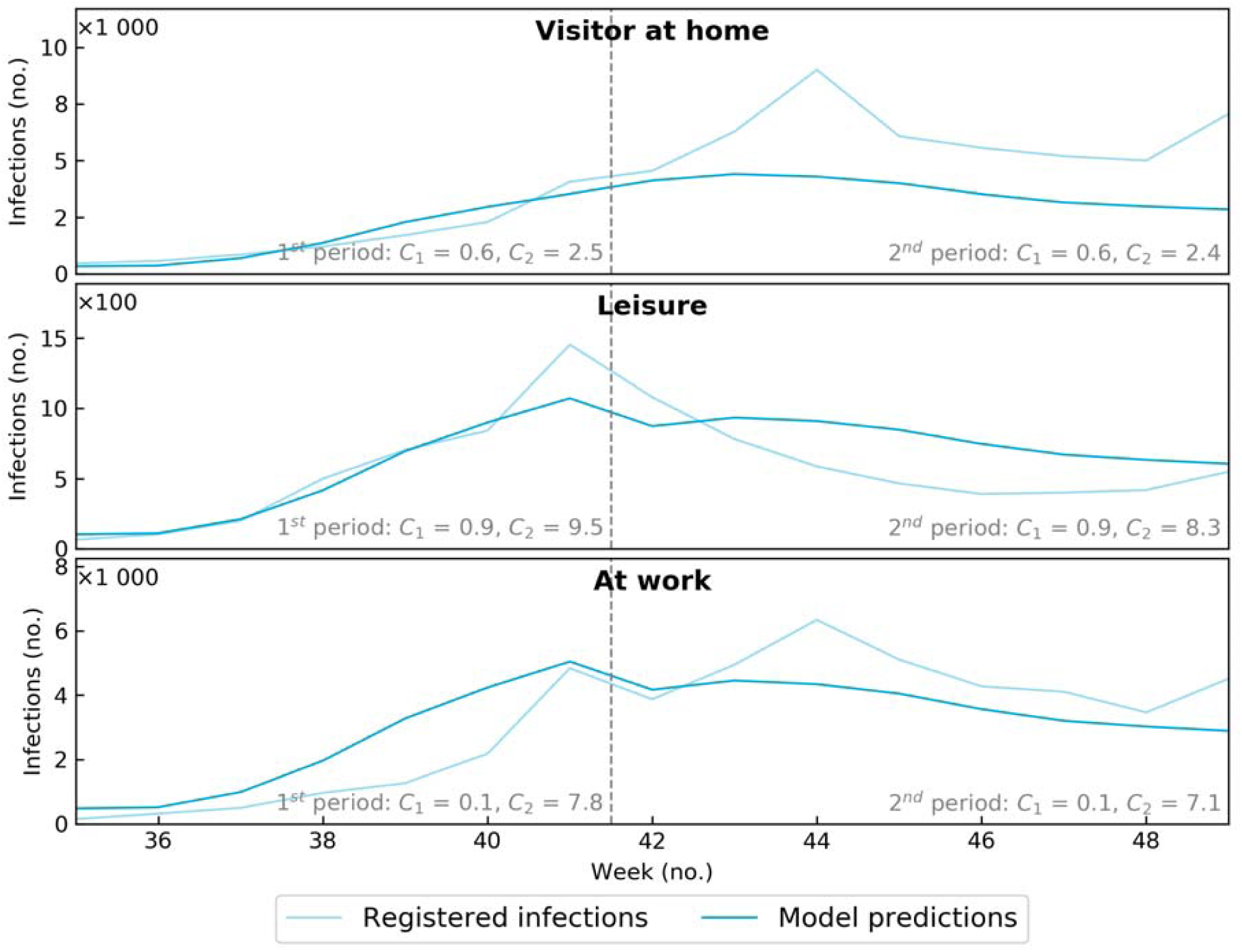
Number of infections based on internal validation by reproduction of infections at work, visitor at home and leisure.

### 2.3.2 External validation: test-events

The Fieldlab test-events are for an external validation. The test events occurred during the pandemic in the middle of a wave of infections. A lockdown was still in place and vaccination was not available yet. The input for the model is defined as:

During these test-events *C*_1_ en *C*_2_ are measured for different variants of measures at these events and different types of events.

All Fieldlab participants and crew were asked to get tested on day five after the event, a request that was followed by more than 80% of the participants. In addition, all positive cases related to a Fieldlab event, identified by the regional Health Care Services, were included in the data set. Infections identified after an event included persons infected just before or after the pre-test or persons who had taken a PCR test around the cut-off of the PCR, thereby varying in outcome. This means that people who were at the event and tested positive also could have been infected at other locations before of after the test-event.

Very short, “passing” contacts of less than 10 seconds were not taken into account because these “passing contacts” are assumed not to be significant with regard to transmission. At the test events, the generic measures for social distancing were not in place. The risk of infection was reduced by several packages comprising variations in occupation rate, catering, crowd management, the use of masks, ventilation protocols.

- *P* is based on the prevalence of the date of the events as published by RIVM (<u>13</u>)
- *V*_1_ = 100 % Because vaccination was not available yet and earlier infections were very limited, so no immunity is taken into account,*F*_*T*,1_ = 0,95. Persons with COVID-19 (like) symptoms were banned from participation. All visitors and crew needed a negative PCR test taken within 48 hours before the event in order to attend. As the PCR test may pick up low viral loads such as in cases of persons who recently recovered from COVID-19, the ratio of positive tests is higher than the ratio of asymptomatic people only (*18*).
- *F*_*y*,0_ = 0 if the ventilation meets the requirements of building codes and *F*_*y*,1_ = 0.90 This value is based on expert judgment. *F*_*y*,1_ is applied when ventilation is significantly improved (with a CO2 value below 800 ppm). Venues have been checked prior to the event and during the event the CO2 value was measured. All outdoor events were not completely open (the festival was in a tent, football stadiums had a large roof), therefore we assumed *F*_*y*,2_ = *F*_*y*,1_.
- The effectivity of masks was estimated by medical experts. While the effectivity under *in-vitro* conditions can be high (*19*), a low estimate is realistic for their effectiveness during *in-vivo* events because masks are not used correctly and they may be ill-fitting. During all test events different rules applied.*F*_*M*_ = 0,05 when the mask was used only when people were seated, and *F*_*M*_ = 0.1 when the mask was used while people move (but not while they are drinking and eating). During type 2 and 4 events, *F*_*M*_ was set at 0, as masks compliance was extremely low to non-existent.

The number (and risk of) infections at the event are estimated with the model based on the prevalence and measures at the test-events. While in the post-event test results, the crew is also taken into account, the calculated risk for infections applies to visitors only. The model results can be compared with the confirmed infections after the events. In Table 4 the results of the test events have been summarized per type of event and compared to the model results. The detailed results of all events and measures are described elsewhere (*20*).

**Table 4:** Comparison model results with realization at test events

|  | Type I | Type II | Type III | Type IV |
| --- | --- | --- | --- | --- |
| People at event ($N$) (without crew) [persons] | 815 | 2341 | 1692 | 2960 |
| Average duration ($t$) of event [hours] | 4.4 | 4.3 | 3 | 7 |
| Average prevalence ($P$) at events | 0.0056 | 0.0061 | 0.0056 | 0.0077 |
| Number of Pretests (incl crew) [persons] | 1198 | 3078 | 2033 | 3890 |
| Positive pretest [persons] | 11 (0.9%) | 18 (0.6%) | 12 (0.6%) | 26 (0.7%) |
| After (5d) tests (incl crew) [persons] | 926 | 2603 | 1689 | 3168 |
| Positive after test[persons] | 1 (0.1%) | 14 (0.5%) | 4 (0.2%) | 26 (0.8%) |
| Possible infections at event (realization) [persons] | 0 | 4 | 0 | 12 |
| Confirmed infections at event (realization) [persons] | 0 | 0 | 0 | 4 |
| Expected Infections ($S$) at event (model) [persons] | 0.04 | 0.28 | 0.05 | 0.54 |
| Expected Infections ($S$) at event without measures (model) [persons] | 0.86 | 5.56 | 1.14 | 10.81 |
| Individual risk ($R_{event}$) per hour at event | $1.12 \cdot 10^{-5}$ | $2.62 \cdot 10^{-5}$ | $1.00 \cdot 10^{-5}$ | $2.61 \cdot 10^{-5}$ |
| Maximum individual risk per hour at event | $1.4 \cdot 10^{-5}$ | $1.5 \cdot 10^{-5}$ | $8.04 \cdot 10^{-6}$ | $1.6 \cdot 10^{-5}$ |
| Minimal individual risk per hour at event | $8.5 \cdot 10^{-6}$ | $4.3 \cdot 10^{-5}$ | $1.6 \cdot 10^{-5}$ | $3.6 \cdot 10^{-5}$ |
| Average individual Risk ($R_{home}$) per hour at home | $1.06 \cdot 10^{-5}$ | $1.14 \cdot 10^{-5}$ | $1.06 \cdot 10^{-5}$ | $1.45 \cdot 10^{-5}$ |
| Average individual Risk ($R_{visitor}$) per hour having a visitor | $4.50 \cdot 10^{-5}$ | $4.82 \cdot 10^{-5}$ | $4.48 \cdot 10^{-5}$ | $6.13 \cdot 10^{-5}$ |
| Individual risk per hour ($R_{event}$) at event without measures | $1.25 \cdot 10^{-4}$ | $5.10 \cdot 10^{-4}$ | $2.24 \cdot 10^{-4}$ | $5.22 \cdot 10^{-4}$ |

In the post-event tests, 14 cases have been identified where persons were possibly infected at the events. Other positive tests were excluded during interviews, for example because participants had known close contacts with SARS-CoV-2 positive cases at their home or events around the same time. But even in the 14 cases where participants were possibly infected, infections could have occurred in other places and situations at any time from around the pre-test, the day of the event, or even a few days after the event. If we use the Dutch average infection risk during the event as a control group we can estimate the number of infections which could be expected outside the event. The time spent at the event represents about 4-10% of the total time where people could be infected. If we also consider the risk at other locations, one or two of the possible infections could be (on average) related to the events.

During the eight test-events, four persons were confirmed to be infected during the event. Confirmed infections are those infections which can be related to each other for example by sequencing or because of proven contacts with other positive tested people during the event. Two infections occurred while travelling home with a contagious person, which caused two infections at the type IV event. These are not infected at the event (and part of the model) but these are related to the event.

As the number of events was limited related to the expected probability distribution, the data analysis would have profited from a higher number of events. Still, the data can be used to check if the model is plausible. We believe that we included nearly all infected people at the event because we combined the information of regular testing procedures and the after-event tests. Overall, we concluded that the outcomes of the test-events support the results of the model.

## 4. Discussion and concluding remarks

### 4.1 Reflection to the validation of the model

In the model, we assumed an average of infectious people in the Netherlands. Our assumptions were actually corroborated by the pre-event test results, which were in the range of the nationally reported prevalence of SARS-CoV-2. However, it might be that the prevalence among the visitors was higher than the assumed average. A first argument is that more young people attended the test events, and these age groups contribute relatively more than elderly (as > 60 years old) to the positive PCR tests (*21*). A second argument is that the test events were held during the lockdown, and the non-risk-averse people who attend these events might also have attended more other activities. On the other hand the susceptibility of younger naïve individuals is less than for elder people (22). Because of these other activities it can be expected that the source of infections for some of the cases identified after the event may be unrelated to the event.

At day 5 after the event, visitors and crew were tested, but our model exclusively calculates the risk for visitors, as contact data of the crew were not measured. During the events, the crew attempted to keep their distance from the participants and wore masks continuously.

In general, we found more PCR-positive people in the pre-event than in the post-event testing. As the PCR test may pick-up low viral loads, some of the people testing positive, especially in the pre-tests group, may have been recovered from COVID-19, consequently resulting in a higher positive test ratio. This in part explains the positivity rate in the pre- and post-event testing. However, in the after test, also people with COVID-19 symptoms were included. Only for (one of) the type 4 events, the ratio of positive tests in the after test was higher than in the pre-test. This corresponds to higher numbers of infections at the event.

A final remark about the model can be made with regard to the data which is used to estimate A_1_ and A_2_. The contacts used to train the model were gathered during a period of a (partial) lockdown. Large scale events were already prohibited or regulated. This could cause an underestimation of the risk in dynamic settings.

Based on the available but scarce information, the current risk taxation model results in plausible results, instead the model cannot be rejected as being invalid. The test events show that the Fieldlab measures at the events, which replace generic social distancing, reduce the risk to a level below the threshold level (which was the risk at home). The model can be extended with the risk for loss of life and hospitalization using the relation with the age of people. However, once more data is available, this larger training set can help improve the model. Once more data of especially type IV and maybe type II events becomes available, the need for a additional factor for increased transmission at dynamic events can be identified and incorporated. We therefore recommend to collect more data about infections and events while the COVID-19 pandemic continues.

### 4.2 Added value for decision makers and event planners

Depending on the package of measures (such as testing, maximum occupation rate, ventilation) and the proportion of infectious people in the society the risk of infection at an event can be reduced. These measures can be an alternative for social distancing at these events which in general will lead to an increase in risk because of the increase in contacts per hour. The test events in the Netherlands showed that the risk at many of these events can be reduced to a level that is equal to the average risk people are exposed to outside these events. This means that events, with supporting measures, can be organized in such a way that they do no contribute more to the risk of infection per hour as other activities which are considered to be more safe. When the total risk for infection in the society however is to high choices can be made which activities have to be stopped. These decisions are outside the scope of our research.

To decide whether an event is acceptable or te decide for the need of additional measures a threshold can be used. In figure 2, such an example is given. The figure shows the expected the risk of infections (per 100.000 people per hour) as a function of the prevalence. Figure 2 holds the same event but with different packages of measures. The horizontal bar is the threshold, now set at 1 or 2 infections per 100.000 people per hour. During the test-events the risk for infection for people at home was about 1 per 100.000 per hour, the average risk of all types locations in the Dutch society was between 1 and 2 per 100.000 per hour. When the risk for an event is above the threshold additional measures are needed as testing, smart design of the catering and crowd control and occupation rates. These measures result in a reduction of the risk, when the risk is below the threshold the risk because of the event is acceptable. The developed COVID-19 Risk taxation method for (large scale) events can be applied to events to define an alternative package of measures replacing generic social distancing. The value for the thresholds is also a political choice. We therefore recommend a debate to discuss them in perspective to an acceptable risk for infection or load to the health care system.

**Figure 2:**
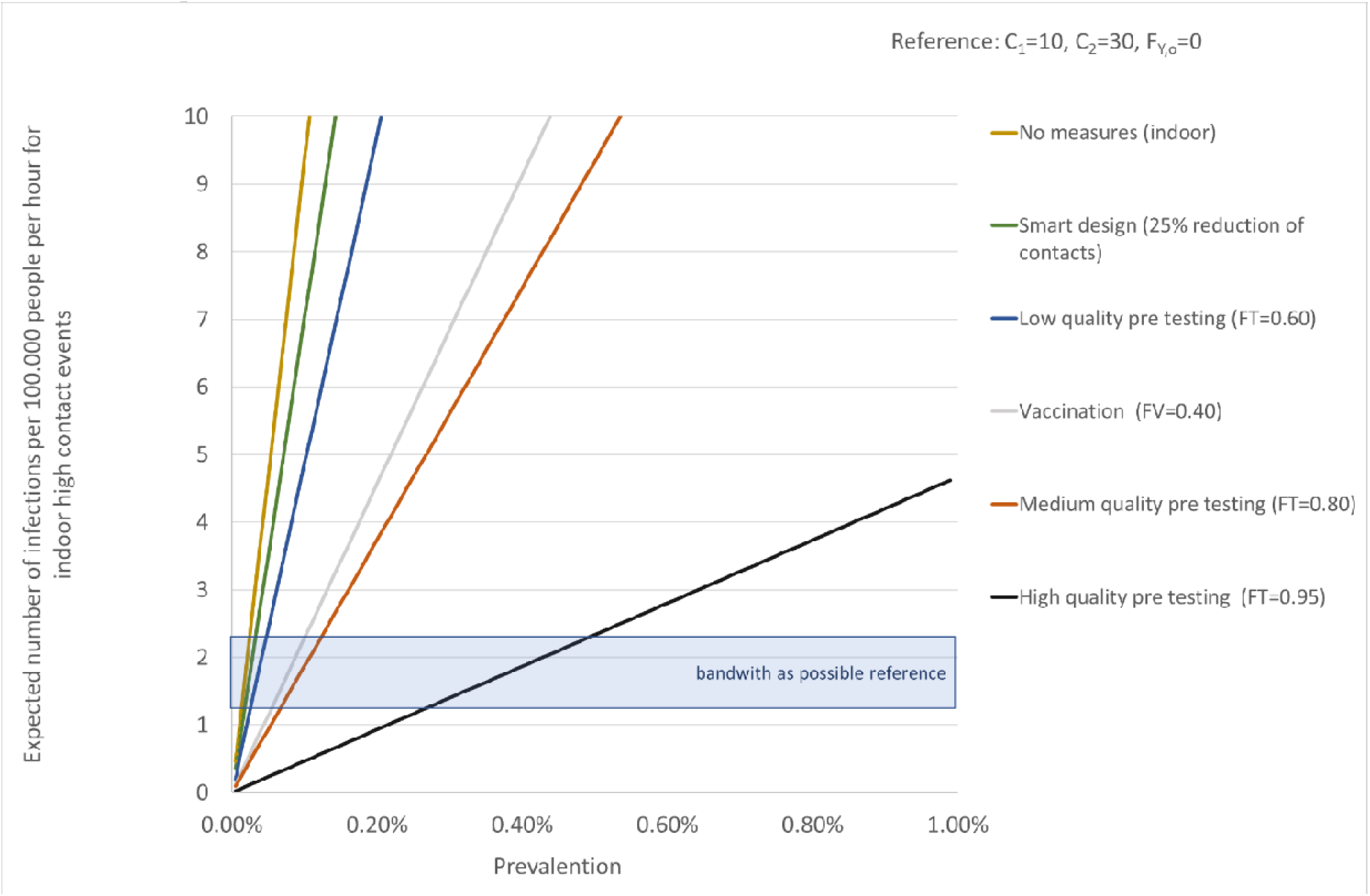
Risk of infections as a function of the prevalence for different packages of measures, related to a possible reference.

## Data Availability

All data produced in the present work are contained in the manuscript

## Notes

### Competing Interest Statement

The authors have declared no competing interest.

### Funding Statement

This study was funded by Click NL, see https://www.clicknl.nl/en/about-clicknl/

### Author Declarations

Human Research Ethics Committee TU Delft (http://hrec.tudelft.nl/) The application is approved by the Ethics Committee on the 4 December 2020. Contact persons: Ir. J.B.J. Groot Kormelink, secretary HREC Dr. Ir. U. Pesch Chair HREC Faculty of Technology, Policy and Management.

### Summary of Updates

We added the reduction of infectiousness and susceptibility because of vaccination and an earlier infection to the model.

